# Metabolic phenotyping reveals a potential link between elevated faecal amino acids, diet and symptom severity in individuals with severe mental illness

**DOI:** 10.1101/2021.12.01.21267133

**Authors:** Jack Jansma, Rogier van Essen, Bartholomeus C.M. Haarman, Anastasia Chrysovalantou Chatziioannou, Jenny Borkent, Magdalini Ioannou, Saskia van Hemert, Iris E.C. Sommer, Sahar El Aidy

## Abstract

The brain-gut axis is increasingly recognized as an important contributing factor in the onset and progression of severe mental illnesses such as schizophrenia spectrum disorders and bipolar disorder. This study investigates associations between levels of faecal metabolites identified using ^1^H-NMR, clinical parameters, and dietary components of forty-two individuals diagnosed in a transdiagnostic approach to have severe mental illness. Faecal levels of the amino acids; alanine, leucine, and valine showed a significant positive correlation with psychiatric symptom severity as well as with dairy intake. Overall, this study proposes a diet-induced link between the brain-gut axis and the severity of psychiatric symptoms, which could be valuable in the design of novel dietary or therapeutic interventions to improve psychiatric symptoms.

## Introduction

While the etiology of schizophrenia spectrum disorders (schizophrenia, schizophreniform disorder or schizoaffective disorder, SSD) and bipolar disorder (BD) is still largely unknown, current hypotheses include genetic, environmental and inflammatory factors (Carvalho et al., 2020; Horrobin et al., 1994; McCutcheon et al., 2020; Müller, 2018) as key contributing factors. Indeed, both disorders have been associated with increased pro-inflammatory status in the blood and brain. Furthermore, deviations of the gut microbiome have been reported in both SSD and BD, highlighting the role of the brain-gut axis (BGA) (Genedi et al., 2019). Importantly, gut-related disorders such as irritable bowel syndrome, are commonly reported in individuals with SSD or BD (Gupta et al., 1997; Lee et al., 2015). Additionally, frequently used medication such as clozapine, olanzapine, haloperidol and lithium have been shown to affect gut motility and microbiota composition (Flowers et al., 2017; Severance et al., 2015; Sublette et al., 2021; Xu et al., 2021), supporting the BGA as a significantly contributing factor in the etiology of these psychiatric disorders. Such findings spiked the interest to better understand the potential role of the gut and its residing microbes, aiming at the development of therapeutic interventions targeting the BGA to ameliorate the severity of psychiatric disorders such as SSD and BD (Ligezka et al., 2020; Lucidi et al., 2021; Mörkl et al., 2020).

Among several existing hypotheses on the origin of SSD and BD, the membrane hypothesis postulates that SSD is a systemic disease originating from increased membrane dysfunction in both the brain and gut (Horrobin et al., 1994), resulting in increased translocation of metabolites, and toxic compounds from the gut to the bloodstream and eventually reaching the brain (Bischoff et al., 2014). A genome-wide association study identified an association between the endothelial cell-selective adhesion molecule (*ESAM*) gene and SSD (Pouget et al., 2016). *ESAM* is expressed on endothelial cells and is associated with tight junction proteins in the blood-brain barrier and the intestinal barrier (Nasdala et al., 2002; Uhlén et al., 2015). In SSD, altered endothelial barrier function in the brain was associated with an influx of macrophages in the brain, increased pro-inflammatory status (Cai et al., 2018) and decreased memory functioning (Cai et al., 2020). Moreover, both SSD and BD individuals have been reported to have increased serum zonulin levels as well as soluble CD14 (Barber et al., 2019; Kılıç et al., 2020; Severance et al., 2013; Usta et al., 2021). Such intestinal barrier dysfunction and the subsequent translocation of (harmful) substances has been closely linked to altered gut microbiota composition (Chelakkot et al., 2018), which was also reported in individuals with BD and SSD (Evans et al., 2017; Nguyen et al., 2019).

To investigate whether gut-associated metabolites are linked with clinical variables, including psychiatric and gastrointestinal symptom severity, we employed untargeted metabolomics using proton nuclear magnetic resonance spectroscopy (^1^H-NMR) on faecal samples collected from forty-two individuals diagnosed in a transdiagnostic approach to have SSD or BD.

## Material and methods

### Participants and clinical variables

As part of an ongoing randomized control study (GUTS, trail register number NL6385), forty-two participants (15 SSD and 27 BD) were analyzed in this baseline cross sectional analysis. The study was approved by the ethics committee of the University Medical Center Groningen (UMCG) (METc 2018/614) and all participants provided written consent upon explanation of the study procedures. To be eligible for participation in the GUTS study, patients were required to have a DSM-IV-R or DSM-5 diagnosis of schizophrenia (295.x), schizophreniform disorder (295.4), schizoaffective disorder (295.7), or bipolar disorder (296.x). Clinical variables including age, sex, body mass index (BMI) and medication intake were reported. Symptom severity was evaluated by trained interviewers using the Brief Psychiatric Rating Scale (BPRS) (Overall and Gorham, 1962) and gastrointestinal complaints were evaluated by the self-report questionnaire: the gastrointestinal symptoms rating scale (GSRS) **(Table 1)** (Kulich et al., 2008). The participants were asked to fill in the Dutch healthy diet index (DHD-index), which is used to score participants according to their adherence to the Dutch guidelines for a healthy diet and measures the nutrient density of their diet **(Table 2)** (Looman et al., 2017; van Lee et al., 2012). Additionally, each subject handed in a faecal sample, which was stored at -80 °C until further use.

**Table 1:**
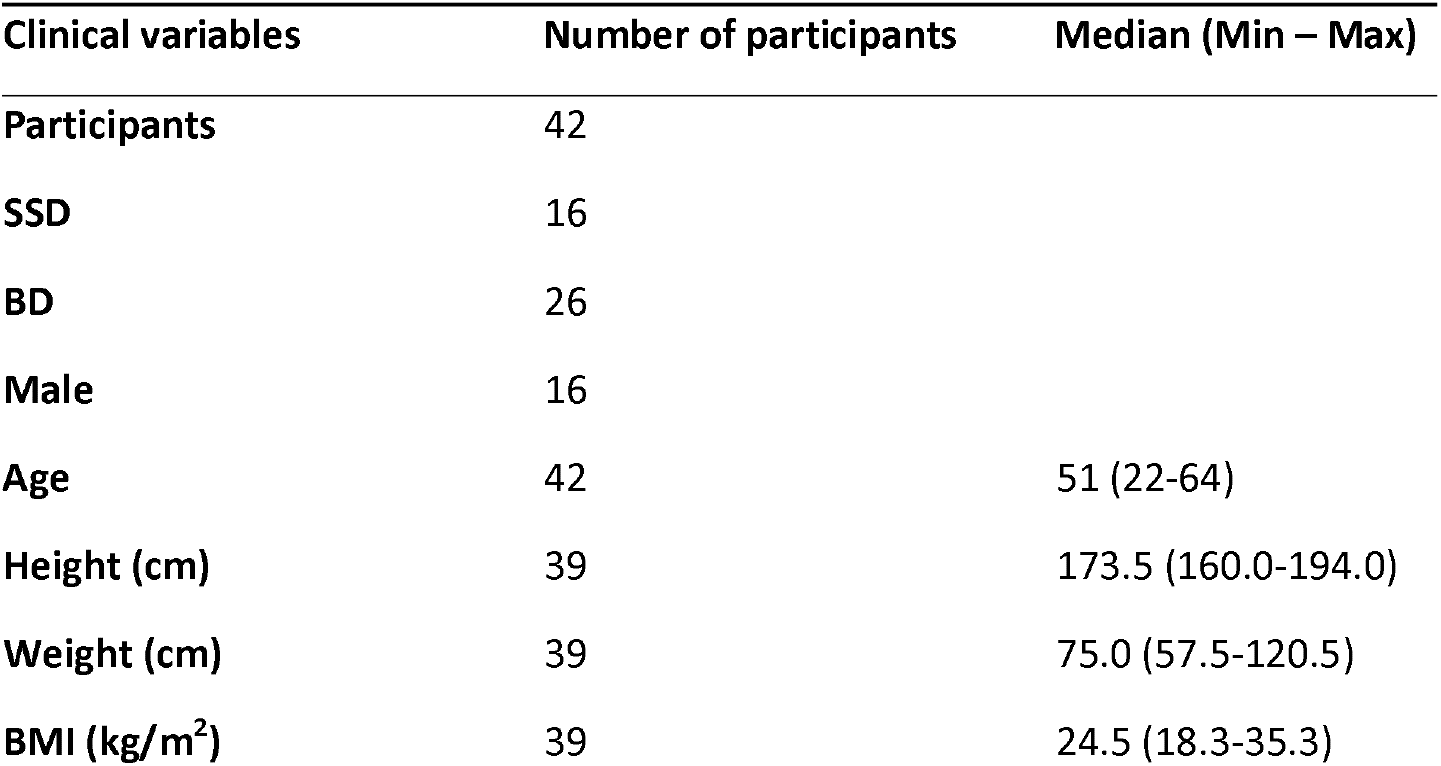
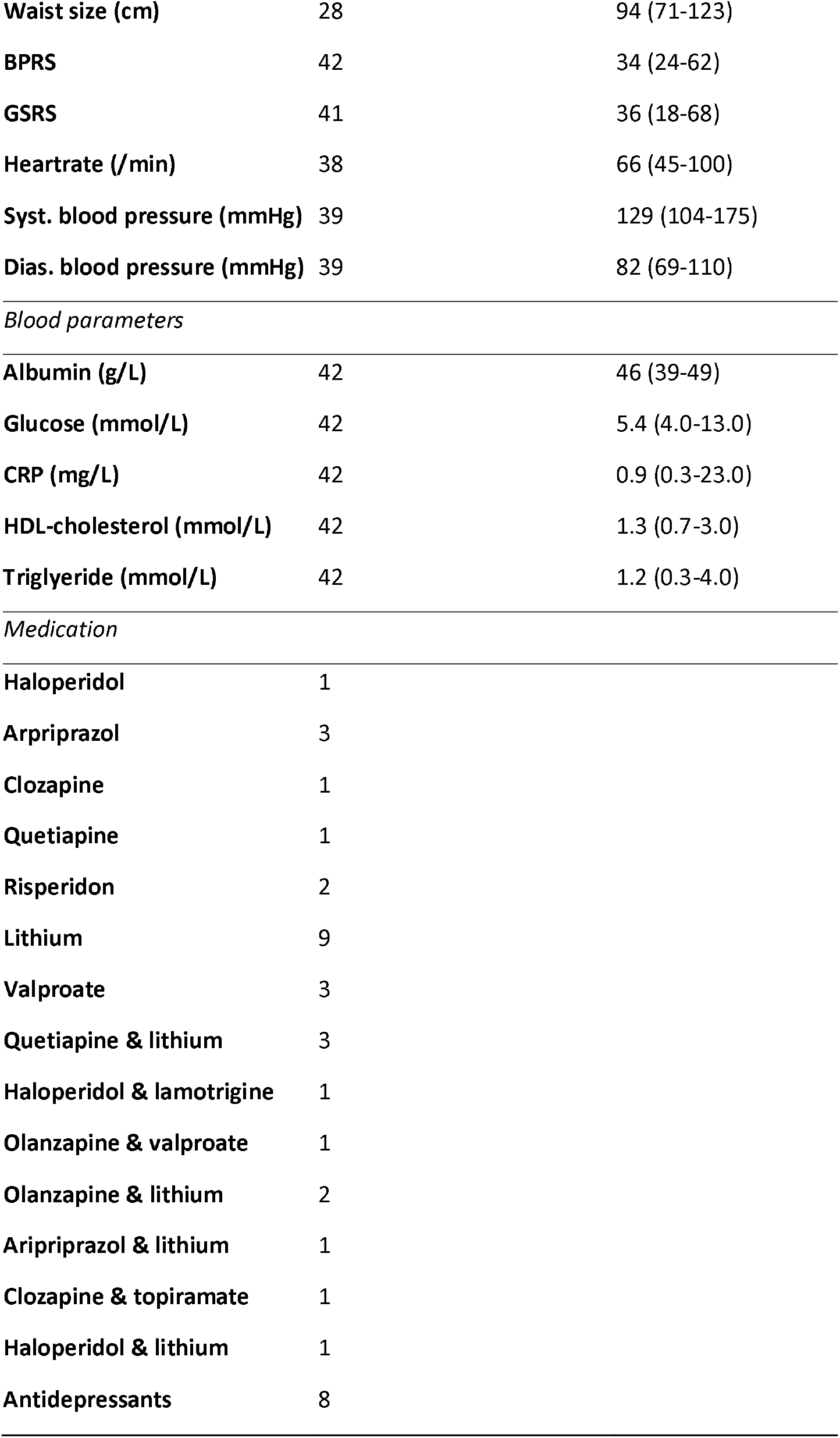
Clinical variables of the cohort. The clinical variables, blood parameters and medication intake of the cohort. The median of each variable is given with the minimal and maximal value in brackets. SSD: schizophrenia spectrum disorder; BD: bipolar disorder; BMI: body mass index; BPRS: brief psychiatric rating scale; GSRS: gastrointestinal symptoms rating scale; Syst: systolic; Dias: diastolic; CRP: C-reactive protein

| Clinical variables | Number of participants | Median (Min – Max) |
| --- | --- | --- |
| Participants | 42 |  |
| SSD | 16 |  |
| BD | 26 |  |
| Male | 16 |  |
| Age | 42 | 51 (22-64) |
| Height (cm) | 39 | 173.5 (160.0-194.0) |
| Weight (cm) | 39 | 75.0 (57.5-120.5) |
| BMI (kg/m<sup>2</sup>) | 39 | 24.5 (18.3-35.3) |
| **Waist size (cm)** | 28 | 94 (71-123) |
| **BPRS** | 42 | 34 (24-62) |
| **GSRS** | 41 | 36 (18-68) |
| **Heartrate (/min)** | 38 | 66 (45-100) |
| **Syst. blood pressure (mmHg)** | 39 | 129 (104-175) |
| **Dias. blood pressure (mmHg)** | 39 | 82 (69-110) |
| *Blood parameters* |  |  |
| **Albumin (g/L)** | 42 | 46 (39-49) |
| **Glucose (mmol/L)** | 42 | 5.4 (4.0-13.0) |
| **CRP (mg/L)** | 42 | 0.9 (0.3-23.0) |
| **HDL-cholesterol (mmol/L)** | 42 | 1.3 (0.7-3.0) |
| **Triglyeride (mmol/L)** | 42 | 1.2 (0.3-4.0) |

*Medication*
|  |  |
| --- | --- |
| **Haloperidol** | 1 |
| **Arpriprazol** | 3 |
| **Clozapine** | 1 |
| **Quetiapine** | 1 |
| **Risperidon** | 2 |
| **Lithium** | 9 |
| **Valproate** | 3 |
| **Quetiapine & lithium** | 3 |
| **Haloperidol & lamotrigine** | 1 |
| **Olanzapine & valproate** | 1 |
| **Olanzapine & lithium** | 2 |
| **Aripriprazol & lithium** | 1 |
| **Clozapine & topiramate** | 1 |
| **Haloperidol & lithium** | 1 |
| **Antidepressants** | 8 |

**Table 2:** Daily intake of participants per category and the macro- and micronutrient density according to the Dutch healthy diet index. The daily intake of the participants per category and the macro- and micronutrient density according to the Dutch healthy diet index (van Lee et al., 2012). The median of each category is given with the minimal and maximal value in brackets. DHD: Dutch healthy diet; Sweet. Bever.: Sweetened beverages; MUFA: monounsaturated fatty acid; PUFA: polyunsaturated fatty acid; PUFA-C3: N3-fatty acids; PUFA-C6: N6-fatty acids; EPA: Eicosapentaenoic acid; DHA: Docosahexaenoic acid.

| DHD index product categories | Number of participants | Median (Min – Max) |
| --- | --- | --- |
| Vegetables (g/day) | 41 | 125.8 (5.4-374.4) |
| Fruit (g/day) | 41 | 138.7 (8.4-305.9) |
| Whole grain products (g/day) | 41 | 139.5 (0-312.6) |
| Refined grain products (g/day) | 41 | 22.6 (0-176.7) |
| Legumes (g/day) | 40 | 8.7 (-0-69.7) |
| Nuts (g/day) | 40 | 11.6 (0-81.2) |
| Dairy without cheese (g/day) | 41 | 133.2 (0-1486.6) |
| Fish (g/day) | 41 | 7.8 (0-19.2) |
| Tea (g/day) | 39 | 150.0 (0-1338.0) |
| Red meat (g/day) | 40 | 9.1 (0-101.5) |
| Processed meat (g/day) | 41 | 8.3 (0-98.3) |
| Sweet. Bever. And fruit juice (g/day) | 41 | 22.1 (0-602.9) |
| Soft fats (g/day) | 41 | 13.0 (0-39.5) |
| **Hard fats (g/day)** | 41 | 0 (0-39.5) |
| **Cheese (g/day)** | 41 | 13.3 (0-40.0) |
| *DHD-index nutrient scores* |  |  |
| **Energy (kJ/day)** | 41 | 6197.16 (3051.52 -12636.45) |
| **Protein (g/day)** | 41 | 51.64 (19.15-141.31) |
| **Vegetable protein (g/day)** | 41 | 27.51 (10.92-53.65) |
| **Total fat (g/day)** | 41 | 64.07 (24.74-107.66) |
| **Saturated fat (g/day)** | 41 | 24.54 (11.89-45.62) |
| **MUFA (g/day)** | 41 | 21.34 (7.72-41.45) |
| **PUFA (g/day)** | 41 | 11.01 (3.08-23.82) |
| **Transfat (g/day)** | 41 | 1.14 (0.40-2.38) |
| **PUFA-C3 (g/day)** | 41 | 1.42 (0.40-2.76) |
| **PUFA-C6 (g/day)** | 41 | 9.29 (2.43-20.45) |
| **Alpha-linolenic acid (g/day)** | 41 | 1.20 (0.39-2.48) |
| **EPA (g/day)** | 41 | 0.06 (0-15) |
| **DHA (g/day)** | 41 | 0.09 (0-0.22) |
| **Cholesterol (g/day)** | 41 | 114.77 (41.01-255.11) |
| **Carbohydrates (g/day)** | 41 | 162.84 (-93.51-369.23) |
| **Monosaccharides (g/day)** | 41 | 59.45 (33.82-209.92) |
| **Polysaccharides (g/day)** | 41 | 95.73 (38.68-209.92) |
| **Fiber (g/day)** | 41 | 19.61 (6.76-35.73) |
| **Water (g/day)** | 41 | 1584.99 (716.56-3146.15) |
| **Alcohol (g/day)** | 41 | 3.13 (0-37.55) |
| **Calcium (mg/day)** | 41 | 601.63 (214.14-2175.02) |
| **Total iron (mg/day)** | 41 | 8.82 (3.46-14.76) |
| **Heme iron (mg/day)** | 41 | 0.37 (0.02-1.94) |
| **Non-heme iron (mg/day)** | 41 | 8.42 (3.38-13.71) |
| **Sodium (mg/day)** | 41 | 1550.17 (716.56-3146.15) |
| **Potassium (mg/day)** | 41 | 2460.72 (1032.51-5778.82) |
| **Magnesium (mg/day)** | 41 | 316.83 (149.82-568.42) |
| **Zinc (mg/day)** | 41 | 7.29 (3.11-15.55) |
| **Selenium (mg/day)** | 41 | 35.38 (9.74-72.12) |
| **Copper (mg/day)** | 41 | 1.18 (0.54-2.21) |
| **Vitamin B1 (mg/day)** | 41 | 0.81 (0.37-1.96) |
| **Vitamin B2 (mg/day)** | 41 | 0.81 (0.32-3.87) |
| **Vitamin B6 (mg/day)** | 41 | 1.22 (0.44-3.88) |
| **Vitamin B12 (µg/day)** | 41 | 2.32 (0.22-7.75) |
| **Vitamin D (µg/day)** | 41 | 2.00 (0.28-6.50) |
| **Vitamin E (mg/day)** | 41 | 8.31 (4.01-18.38) |
| **Vitamin C (mg/day)** | 41 | 70.53 (15.96-190.76) |
| **Retinol (µg/day)** | 41 | 317.30 (117.77-824.17) |
| **Folic acid equivalents (µg/day)** | 41 | 196.81 (93.28-512.60) |
| **Folic acid (µg/day)** | 41 | 189.38 (93.29-416.75) |

### Lyophilization and metabolite extraction

To enable comparison of dry weight and homogenization, the faecal samples were lyophilized. Samples were transferred to a precooled glass vial. The samples were lyophilized overnight in a Zirbus VaCo 5/II freeze dryer at -80 °C, with a vacuum of 0.10 mbar. For metabolite extraction, 20 mg of lyophilized sample was used. A volume of 600 μL buffer 1 (D_2_O, 3 mM NaN_3_ and 1 mM TSP, pH 7.4) and 600 μL buffer 2 (200 μL methanol-d4 and 400 μL chloroform-d) was added to the lyophilized faecal material. The samples were vortexed and incubated for 10 minutes at 4 °C. The samples were then centrifuged for 10 minutes, 1100 g at 4 °C. Afterwards, 600 μL of the aqueous phase was transferred to an 5 mm NMR tube. Preliminary experiments comparing the ^1^H-NMR spectra of the aqueous and organic phase showed improved signal in the aqueous phase compared to the organic phase due to removal of the lipid background as described by Hauser et al. (Hauser et al., 2019).

### ^1^H NMR spectroscopy

All NMR spectra were recorded using a Bruker 600 MHz AVANCE II spectrometer equipped with a 5 mm triple resonance inverse cryoprobe and a z-gradient system. The temperature of the samples was controlled at 25 °C during measurement. Prior to data acquisition, tuning and matching of the probe head followed by shimming and proton pulse calibration were performed automatically for each sample. One-dimensional (1D) ^1^H NMR spectra were recorded using the first increment of a NOESY pulse sequence with presaturation (*γ*B_1_ = 50 Hz) for water suppression during a relaxation delay of 4 s and a mixing time of 10 ms. 64 scans of 65,536 points covering 12,335 Hz were recorded and zero filled to 65,536 complex points prior to Fourier transformation, an exponential window function was applied with a line-broadening factor of 1.0 Hz.

### ^1^H NMR data processing

The spectra were phase and baseline corrected and referenced to the internal standard (TSP; *δ* 0.0 ppm), using the MestReNova software (v.12.0.0-20080, Mesterlab Research). With the same software, spectral alignment was performed selecting manually areas and applying a linear filling method of the missing values, after the exclusion of the water signal and the surrounding empty area (4.70-5.16 ppm). Spectral binning followed from -0.50 to 9.00 ppm with an equal size binning step of 0.001 ppm. Noise removal was performed by averaging each integrated bin separately, and removing the bins with an average below 0.144. The annotation of the bins, highlighted as significant by the statistical analysis, was performed with the Chenomx Profiler software (Chenomx NMR Suite 8.6 and Chenomx 600 MHz, version 11) and the HMDB database 5.0 (http://www.hmdb.ca).

### Statistical analysis

MetaboAnalyst (version 5.0, https://www.metaboanalyst.ca/) was used for multivariate statistical analysis of the ^1^H-NMR spectra (Pang et al., 2021). Pearson’s correlation analysis combined with the Bonferroni correction method (α<0.05) was performed to correlate the bins with the patient data. For confirmation of the obtained associations between the amino acids and the clinical variables as well as the associations between the amino acids and the DHD-index, Pearson’s correlation combined with the Benjamini-Hochberg correction method (α<0.05) was performed. All statistical analysis was performed using the psych and corrplot packages in R software version 4.0.4.

## Results

### Faecal amino acids correlate with the severity of psychiatric symptoms

Forty-two participants (15 SSD and 27 BD), diagnosed with either SSD or BD were analyzed in a baseline cross sectional analysis as described in the methods section. To explore the potential differences in the metabolic profiles among the recruited individuals, faecal samples were analyzed using ^1^H-NMR. The ^1^H-NMR spectral data **(Figure 1A)** were binned and subjected to principal component analysis (PCA). No distinction among the samples obtained from the patients could be observed **(Figure 1B)**. Accordingly, the samples were not separated into BD and SSD subgroups in further analysis.

**Figure 1:**
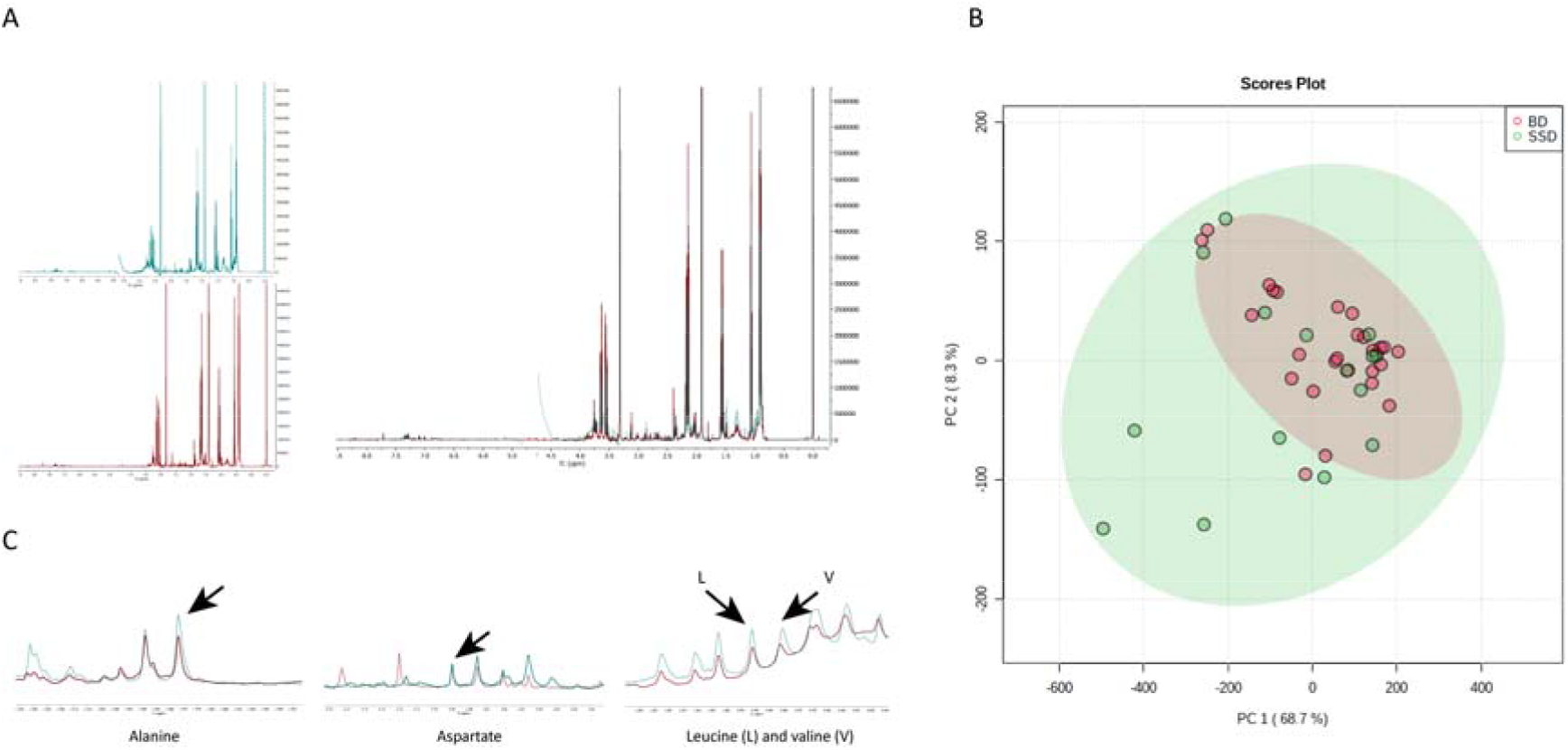
Representative examples of ^1^H-NMR spectra. A) Two ^1^H-NMR spectra representative for an individual with BD and a low (red) and an individual with SSD and a high (green) BPRS score. Both spectra are shown separately and overlapping. The area of water (4.70 – 5.16 ppm) was removed. B) The scores plot of principal component analysis (PCA) based on the first two principal components (PCs), which explain 77.0 % of the variance (68.7 % and 8.3 % for PC1 and PC2 respectively). C) Two ^1^H-NMR spectra representative for an individual with BD and a low (red) and an individual with SSD and a high (green) BPRS score. The integrated peaks are indicated with black arrows. SSD: schizophrenia spectrum disorder; BD: bipolar disorder.

The spectral bins were further processed, and the bins were correlated using Pearson correlation with all the clinical characteristics of the patients **(Table 1)**. Among the correlated bins, 76 significantly correlated with the brief psychiatric rating scale (BPRS) (Overall and Gorham, 1962). Using the library of Chenomx profiler software and the online human metabolome database version 5.0, 51/76 significantly correlated bins were assigned to 4 key-metabolites: alanine, aspartate, leucine, and valine. To confirm the positive association between the identified segments and the BPRS, a representative peak in the ^1^H-NMR spectrum **(Figure 1C)** for each amino acid was integrated, normalized to trimethylsilylpropanoic acid (TSP), and correlated with the BPRS using Pearson correlation **(Figure 2)**. The correlations confirmed the positive association found between alanine (*r* = 0.64, p value = 4.9e-5), leucine (*r* = 0.64, p value = 4.9e-5) and valine (*r* = 0.61, P = 1.4e-4) and the BPRS, respectively. No other correlations were observed between the identified three amino acids and any other clinical parameter, implying a possible role of the increased faecal levels of alanine, leucine and valine association with higher psychiatric symptom severity **(Figure 3)**.

**Figure 2:**
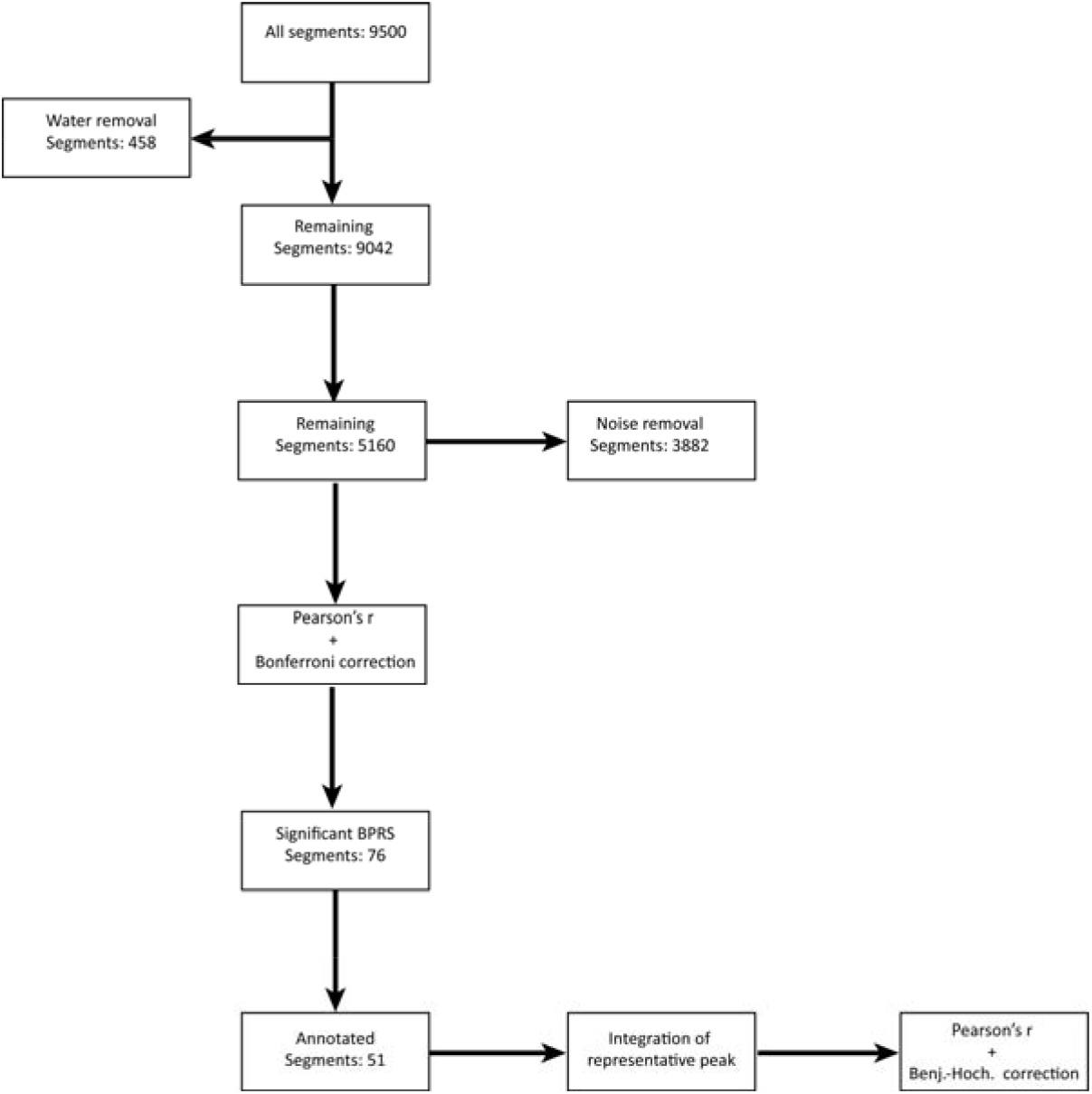
Schematic overview of the workflow starting with the obtained ^1^sH-NMR spectra. A schematic overview depicting the workflow performed starting with the binned spectra. For each step, the number of bins removed or retained is indicated. BPRS: brief psychiatric symptom scale; Benj.-Hoch.: Benjamini-Hochberg.

**Figure 3:**
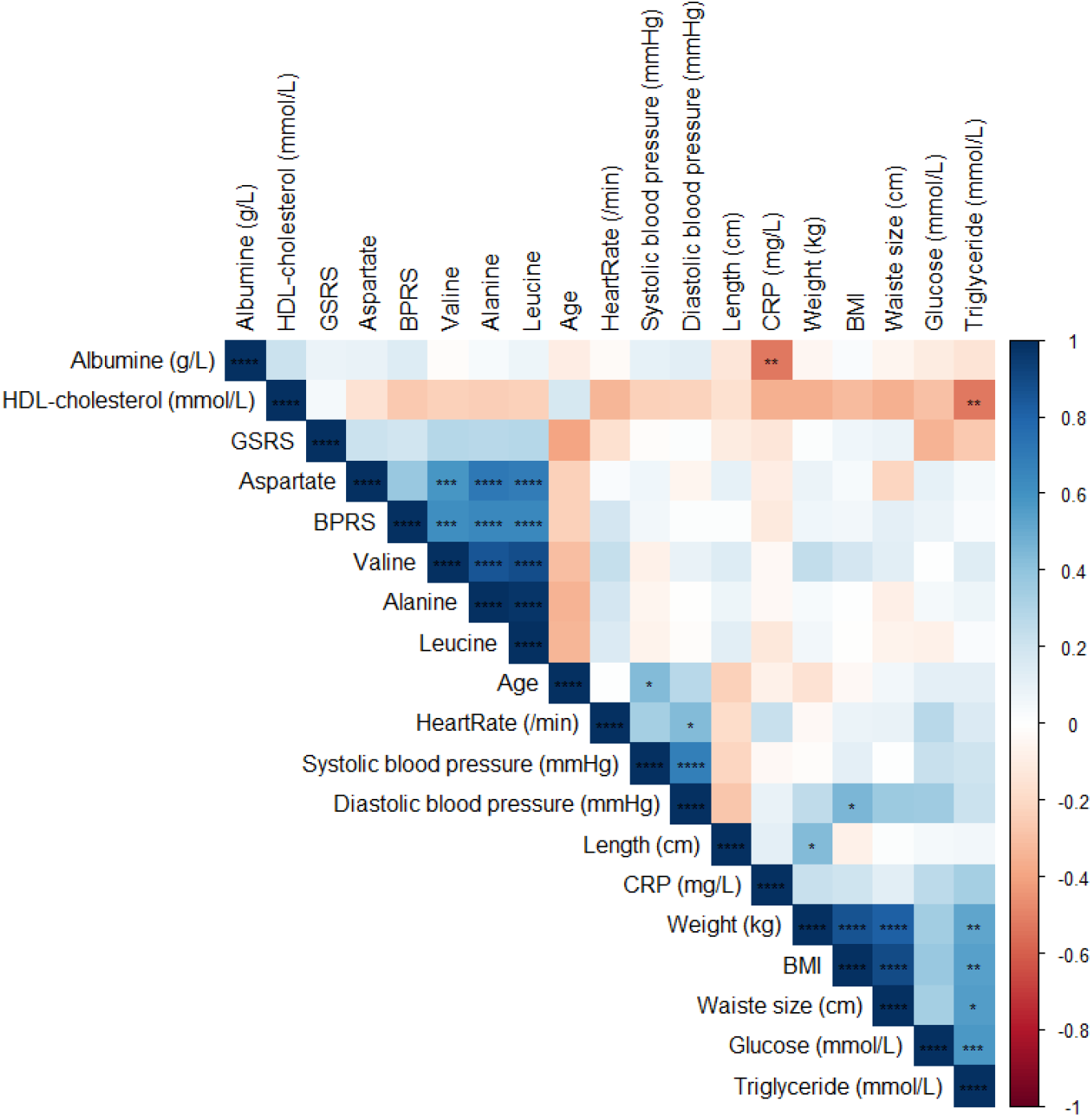
Pearson’s correlation of the clinical variables and amino acids. Pearson’s r correlation between the relative area under the curve of a representative peak corresponding to the amino acids and clinical variables. *, *P* <0.05, **, *P*<0.01, ***, *P*<0.001, ****, *P*<0.0001. GSRS: gastrointestinal rating scale; BPRS: brief psychiatric symptom scale; CRP: C-reactive protein; BMI: body mass index.

Next, we observed strong correlations among the levels of the faecal alanine, leucine and valine; valine-leucine (*r* = 0.89, p value = 8.12e-14); valine-alanine (*r* = 0.86, p value = 6.62e-12); leucine-alanine (*r* = 0.98, p value = 1.14e-27), inferring a potential common source of these amino acids. To this end, the nutrient scores obtained by the Dutch healthy diet index (DHD-index) (van Lee et al., 2012) **(Table 2, 3)** were correlated using Pearson’s correlation, with the faecal amino acid levels. Indeed, positive associations between the total dairy intake without cheese, faecal alanine (*r* = 0.64, p value = 3.9e-4) and leucine (*r* = 0.59, p value = 1.14e-3) were observed **(Figure 4A)**. Moreover, the macro- and micronutrients typically present in high levels in dairy products such as calcium, potassium, vitamin B2, vitamin B6 and vitamin B12 (Pereira, 2014) were positively associated with the faecal levels of alanine (calcium: *r =* 0.5, p value = 0.012; potassium: *r* = 0.48, p value = 0.015; vitamin B2: *r* = 0.63, p value = 7.2e-4; vitamin B6: *r* = 0.63, p value = 7.2e-4; vitamin B12: *r* = 0.48, p value = 0.015) and leucine (calcium: *r =* 0.48, p value = 0.015; potassium: *r* = 0.49, p value = 0.013; vitamin B2: *r* = 0.6, p value = 9.4e-4; vitamin B6: *r* = 0.6, p value = 9.4e-4; vitamin B12: *r* = 0.45, p value = 0.02). The caloric and protein intake showed a positive association with the faecal amino acids alanine (*r* = 0.43, p value = 0.032; *r* = 0.54, p value = 0.0047) and leucine (*r =* 0.44, p value = 0.029; *r* = 0.54, p value = 0.000047) **(Figure 4B)**. Taken together our data show significant positive associations between the dairy-related amino acids detected in faecal samples of trans-diagnosed individuals and the severity of disease symptoms.

**Figure 4:**
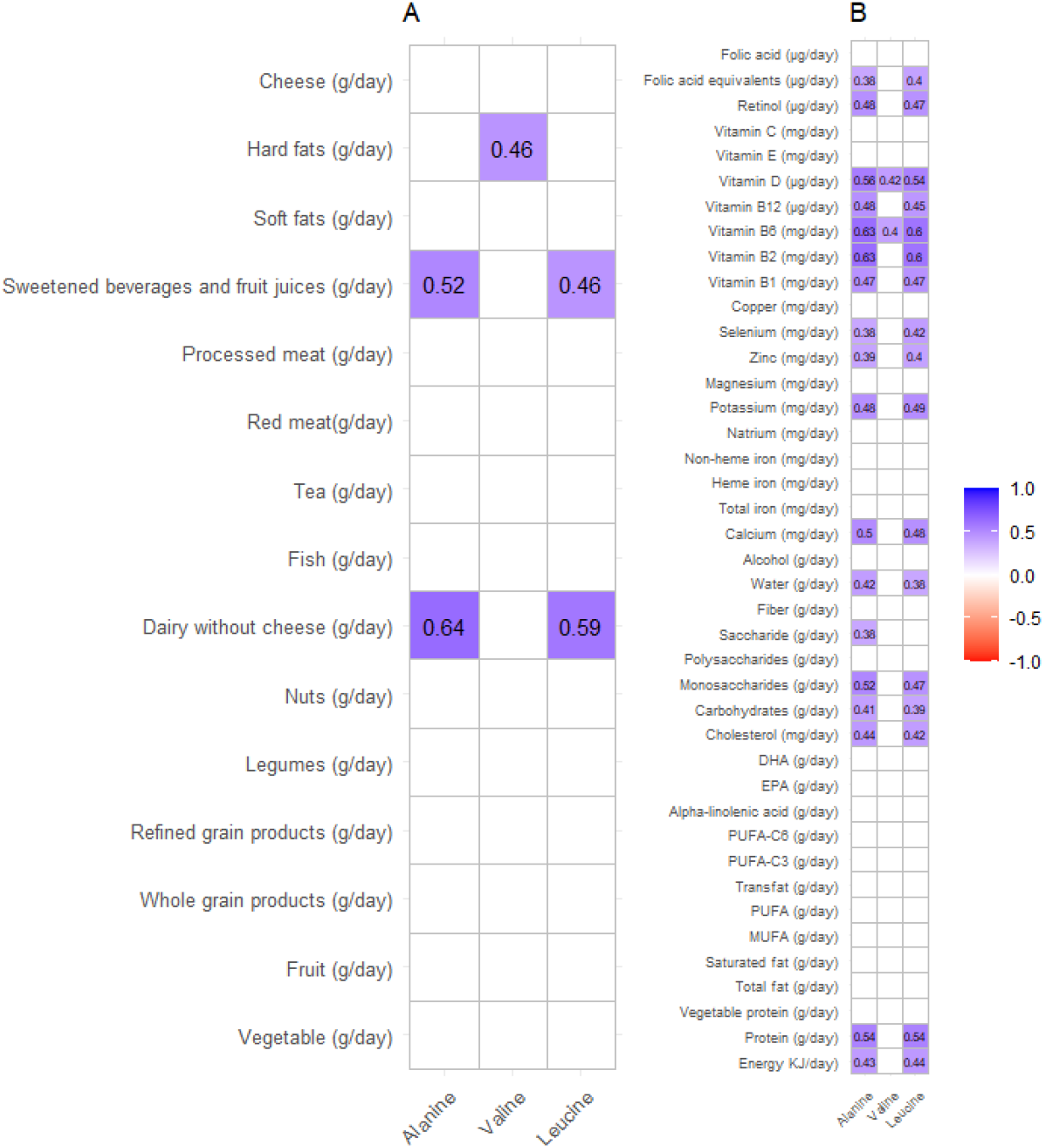
Pearson’s correlation of the amino acids and the BPRS with the Dutch healthy diet index. A) Pearson’s correlations between the daily intake of participants per category according to the Dutch healthy diet (DHD) index (van Lee et al., 2012). B) Pearson’s correlations between the daily macro and micro nutrient intake of participants according to the DHD index. Only significant correlations are colored (P<0.05). The correlation coefficient is given for each significant association.

## Discussion

In this study, ^1^H-NMR was employed to investigate a potential role of intestinal metabolites in mental illness in a trans-diagnostic approach of forty-two individuals with BD or SSD. PCA analysis confirmed the similarity of pathophysiological processes with regard to faecal metabolites among individuals with BD or SSD, as no separation of the metabolic profile between the two groups was detected **(Figure 1B)**.

The positive association reported in this study between the faecal amino acids, alanine, leucine, valine and the psychiatric symptom severity is in agreement with other metabolomics studies performed on serum, urine and cerebrospinal fluid from patients with BD. For example, elevated levels of alanine were reported in BD individuals compared to healthy controls (Sethi et al., 2017a; Xu et al., 2014; Yoshimi et al., 2016). Analogously, higher levels of valine were detected in urine and serum samples of SSD individuals, respectively (Yang et al., 2011).

Our findings point to a possible common source, dairy products, of the three amino acids elevated in the faecal samples of the patients. In fact, Severance et al. showed a positive correlation between the negative psychiatric symptoms according to the positive and negative symptom scale (PANSS) in SSD patients and serum levels of immunoglobulin G (IgG) against the α_s_ and κ subunit of bovine casein, the major protein component in milk (Severance et al., 2010). Interestingly, leucine is the major amino acid in bovine casein (Rafiq et al., 2016). Other studies have found increased levels of serum immunoglobulin A and IgG against gliadin, one of the major components of gluten, in individuals with BD or SSD compared to healthy controls, though the levels did not correlate with psychiatric symptom severity according to the PANSS (Dickerson et al., 2011, 2010). Besides, countries with a high national intake of sugar and dairy products have been reported to have a more severe 2-year outcome of SSD (Peet, 2004), highlighting a key role of the diet in the severity of mental illnesses.

In addition to the diet being a plausible source of the metabolites we detected; the increased levels of these amino acids may have originated from the metabolization of the dietary components by the gut microbiota. Indeed, Zheng et al. showed an increase of, among others, alanine, leucine, and valine in the serum and faecal samples of mice transplanted with faecal material from SSD individuals. The microbiota-based altered metabolites were further detected in the hippocampus of mice transplanted with SSD faecal material and were accompanied by behavioral changes related to SSD (Zheng et al., 2019), highlighting a significant role of the gut microbiota and their metabolic capacity in the development of SSD.

The potential adverse (indirect) effects of the diet are possibly mediated by an impaired intestinal barrier function. This results in increased translocation of gut- and microbiota related metabolites from the lumen of the gut to the bloodstream and eventually the brain, accompanied by an increased inflammatory response (Bischoff et al., 2014). Recently, Carloni et al. showed that inflammation originating in the gut can induce propagation of the inflammation to the brain via increasing permeability of the blood-brain barrier, which may be responsible for the observed mental symptoms in individuals with inflammatory bowel disease (Carloni et al., 2021). Indeed, abnormal immune responses have been reported both in patients with SSD and BD, of varying disease stages and medication status (Beumer et al., 2012; Horváth and Mirnics, 2014; Muller and J. Schwarz, 2010; Munkholm et al., 2013). In systematic reviews involving thousands of patients, both disorders were robustly associated with increased C-reactive protein (CRP) levels, especially during early-phase acute psychosis (Fond et al., 2018) and during mania (Fernandes et al., 2016). Moreover, both disorders have been reported to be associated with an increased pro-inflammatory gene expression in circulating monocytes (Drexhage et al., 2010) and symptom severity was found to correlate with levels of inflammatory markers (Fan et al., 2010, 2007; Hope et al., 2013). Similarly, a meta-analysis comparing cytokine profiles in patients with SSD, BD, and major depressive disorder demonstrated manifest alterations in blood cytokine levels most consistent with an inflammatory profile and T-cell activation (Goldsmith et al., 2016). This analysis with high scientific impact concluded that despite between-study heterogeneity, there were remarkable similarities in the patterns that suggest that abnormal immune responses may be, at least partially, responsible for the reported findings. Taken together, our results highlight a potential link between elevated faecal amino acids, diet and symptom severity in individuals with severe mental illness and urges for the need for further investigations into the interplay between diet, brain and gut.

### Limitations

The results of this research should be interpreted in light of some limitations. The small sample size including potential medication effect precludes the conclusion of any causation and limits the correlation analysis, as well as the lack of a matched control group. Moreover, although ^1^H-NMR spectroscopy-based metabolomics analysis has been shown to be a useful strategy to investigate the pathophysiology of BD and SSD (Sethi et al., 2017b; Tasic et al., 2017), particularly in serum samples, the sensitivity of ^1^H-NMR is relatively low. Therefore, metabolites in concentrations below the detection limit are missed, which impacts the outcome of the study. Nonetheless, we consider this pioneer work and encourage more extensive research into the BGA in SSD and BD.

### Conclusion

Overall, the present study shows a significant role of the interplay between the diet-brain-gut axis in the severity of mental illness, which helps guide the development of novel dietary, and microbiota-based interventions.

## Data Availability

The data presented in this study are available on request from the corresponding author

## Author Contributions

Jack Jansma: Formal analysis, writing-original draft. Rogier van Essen: Conceptualization, methodology, investigation. Bartholomeus C.M. Haarman: Conceptualization, writing-review and editing, funding acquisition. Anastasia Chrysovalantou Chatziioannou: methodology, writing-review and editing. Jenny Borkent: writing-review and editing. Magdalini Ioannou: writing-review and editing. Saskia van Hemert: Conceptualization. Iris E.C. Sommer: Conceptualization, funding acquisition, writing-review and editing. Sahar El Aidy: Conceptualization, writing-review and editing, funding acquisition. All authors have read and agreed to the published version of the manuscript.

## Institutional Review Board Statement

The study was conducted according to the guidelines of the Declaration of Helsinki, and approved by the Institutional Review Board (or Ethics Committee) of University Medical Center Groningen (UMCG) (METc 2018/614; March 19, 2019).

## Informed Consent Statement

Informed consent was obtained from all subjects involved in the study.

## Data Availability Statement

The data presented in this study are available on request from the corresponding author, upon completion of the study.

## Acknowledgements

We thank Dr. Johan Kemmink and ing. Pieter van der Meulen of the Stratingh institute of chemistry, University of Groningen, The Netherlands for assisting us with the ^1^H-NMR measurements.

## List of abbreviations

BD: Bipolar disorder
BGA: Brain-gut axis
BMI: Body mass index
BPRS: Brief psychiatric rating scale
CRP: C-reactive protein
DHD-index: Dutch healthy diet index
ESAM: Endothelial cell-selective adhesion molecule
GSRS: Gastrointestinal symptoms rating scale
^1^H-NMR: Proton nuclear magnetic resonance
IgG: Immunoglobulin G
PANSS: Positive and negative symptom scale
SSD: Schizophrenia spectrum disorders

